# Effectiveness of Vaccination against Reported SARS-CoV-2 Infection in United States Coast Guard Personnel between May and August 2021: A Time-Series Analysis

**DOI:** 10.1101/2021.11.19.21266537

**Authors:** John Iskander, Jamie Frost, Sharon Russell, Jaspal Ahluwalia, Emily Ward, Shane Steiner, Dana Thomas, Paul Michaud

## Abstract

**Background:** The United States Coast Guard (CG) began voluntary use of SARS-CoV-2 vaccines under an Emergency Use Authorization on December 16, 2020. Vaccination status is monitored through a service-wide immunization registry. Active Duty and Reserve (military) CG members are required to report any new positive test for COVID-19 to a centralized database.

**Methods:** Between May and August 2021, vaccination effectiveness (VE) against any new report of COVID-19 was calculated according to standard formulas, using registry immunization status of cases and monthly mid-point vaccine coverage data. CG members recorded as fully vaccinated with a two-dose vaccine were compared with those with any other vaccine status. Sub-analyses were also conducted according to geographic area (Atlantic vs Pacific), age, and type of vaccine received.

**Results:** Effectiveness of full vaccination reached a peak of 89.0% in June, then declined over the rest of the study period to 62.7% in August. In July and August, steeper declines in VE were seen in the Atlantic region. The rate of breakthrough infections remained under 1% in two-dose vaccine recipients, and did not differ between those who received the Moderna or Pfizer vaccines. No hospitalizations or deaths due to COVID-19 disease were recorded in fully vaccinated Coast Guard members.

**Conclusions:** Coincident with the national spread of the Delta variant of SARS-CoV-2, overall vaccine effectiveness among CG personnel decreased during the summer months of 2021, but continued to provide substantial protection, as well as full protection against the most serious outcomes. Policy initiatives and outreach intended to increase vaccine coverage within this and other military populations could extend the disease prevention benefits seen in this study.

## Introduction

Infection with SARS-CoV-2, the virus that causes COVID-19 disease, threatens the health of individual military service members as well as military operational readiness more generally. Service members may have occupational risk factors for acquiring SARS-CoV-2, including working and living in close congregate settings (Marcus JE et al 2020) and being deployed in support of COVID-19 related missions (Clifton GT et al, 2021). Additionally, military personnel are predominantly young adults who are more likely to have asymptomatic infections (Yonker et al., 2020).

Following the U.S. Food and Drug Administration (FDA) granting of Emergency Use Authorization (EUA) for mRNA-based COVID-19 vaccines produced by Pfizer and Moderna, in December 2020 the United States Coast Guard (CG) began vaccinating its military (Active Duty and Reserve) and civilian members on a voluntary basis. While there have been reports of post-authorization COVID-19 vaccine effectiveness (VE), there have been no published studies of VE in military populations; recent analyses have noted changing patterns of VE associated with wider spread of the Delta variant of SARS-CoV-2 (Rosenberg ES et al 2021, Herlihy R et al 2021, Lopez Bernal J et al 2021). In order to monitor recent trends in VE and to describe the performance of vaccines within our service, we used COVID-19 disease reporting and vaccine monitoring systems maintained within the CG to conduct sequential time-series analyses of VE.

## Methods

### Case Ascertainment

The Coast Guard Personnel Accountability and Assessment System (CGPAAS) provides continuously updated data on individuals reporting positive tests for COVID-19. Since June 2020, military personnel with access to the system are required to report any new positive test; civilian reporting is voluntary. For the study months of May through August 2021, we assigned cases to either the date the CG member first entered their report into the system, or the date they reported that their test was positive, whichever was earlier. To allow for reporting delays, reports received through September 15 2021 were included in the analysis as part of August data if they reported a diagnosis date before the end of August. For the study months, using a pre-existing hospital discharge data set we also monitored all COVID-19 related hospitalizations among CG personnel.

### Vaccination Status Ascertainment

We assessed vaccination status using a CG wide immunization registry which tracks the status of ∼42,000 Active Duty, ∼6,300 Reserve, and ∼8,500 civilian personnel (CG, unpublished data). Civilians are not required to report immunizations, but may do so voluntarily. Data is updated multiple times per day. For monthly analyses, we used data on number of persons fully vaccinated (defined as having received two doses of vaccine) and in all other vaccination categories from the 16^th^ for months with 31 days, and from the 15^th^ for months with 30 days. Vaccination status of personnel reporting a new COVID-19 diagnosis or hospitalized due to COVID-19 was assessed using a unique identifier which linked the registry with CGPAAS or hospitalization data.

### Calculation of Vaccine Effectiveness and Sub-analyses

Because of evidence of limited effectiveness of a single dose of mRNA vaccine against the Delta variant (Lopez Bernal J et al, 2021) and in accordance with other analyses (Rubin D et al, 2021), for our primary analysis we dichotomized the data into those who were fully vaccinated compared to all others. Because >99 percent of vaccine used within CG was either the Pfizer or Moderna 2-dose mRNA based vaccines (data not shown), we excluded from our primary VE analysis those who had received the single dose Johnson and Johnson (J and J) vaccine.

We used the following standard formula (Orenstein W et al, 1985) to calculate vaccine effectiveness:

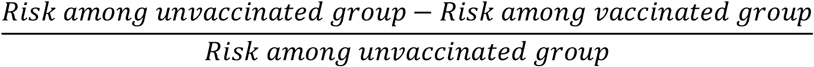

For comparability with other studies we transformed this calculation into percentages, and calculated 95% confidence intervals (CI) using published formulas (Orenstein W et al, 1985). For each month, we measured the risk among not fully vaccinated persons reporting a new positive test for COVID-19 divided by the midpoint month number of persons recorded as other than fully vaccinated. Risk among fully vaccinated CG personnel was calculated in a parallel manner. Using the same formula, we also calculated VE for Atlantic (LANT) and Pacific (PAC) regions (Figure 1), and by pre-specified age categories (17-24 y/o, 25-31 y/o, 32-44 y/o, and > 45 y/o). Because preliminary analysis indicated that the vast majority of COVID-19 reports to CGPAAS were from military personnel and that overall VE did not differ between military and civilian personnel, we did not disaggregate the data by military vs civilian status.

**Figure 1:**
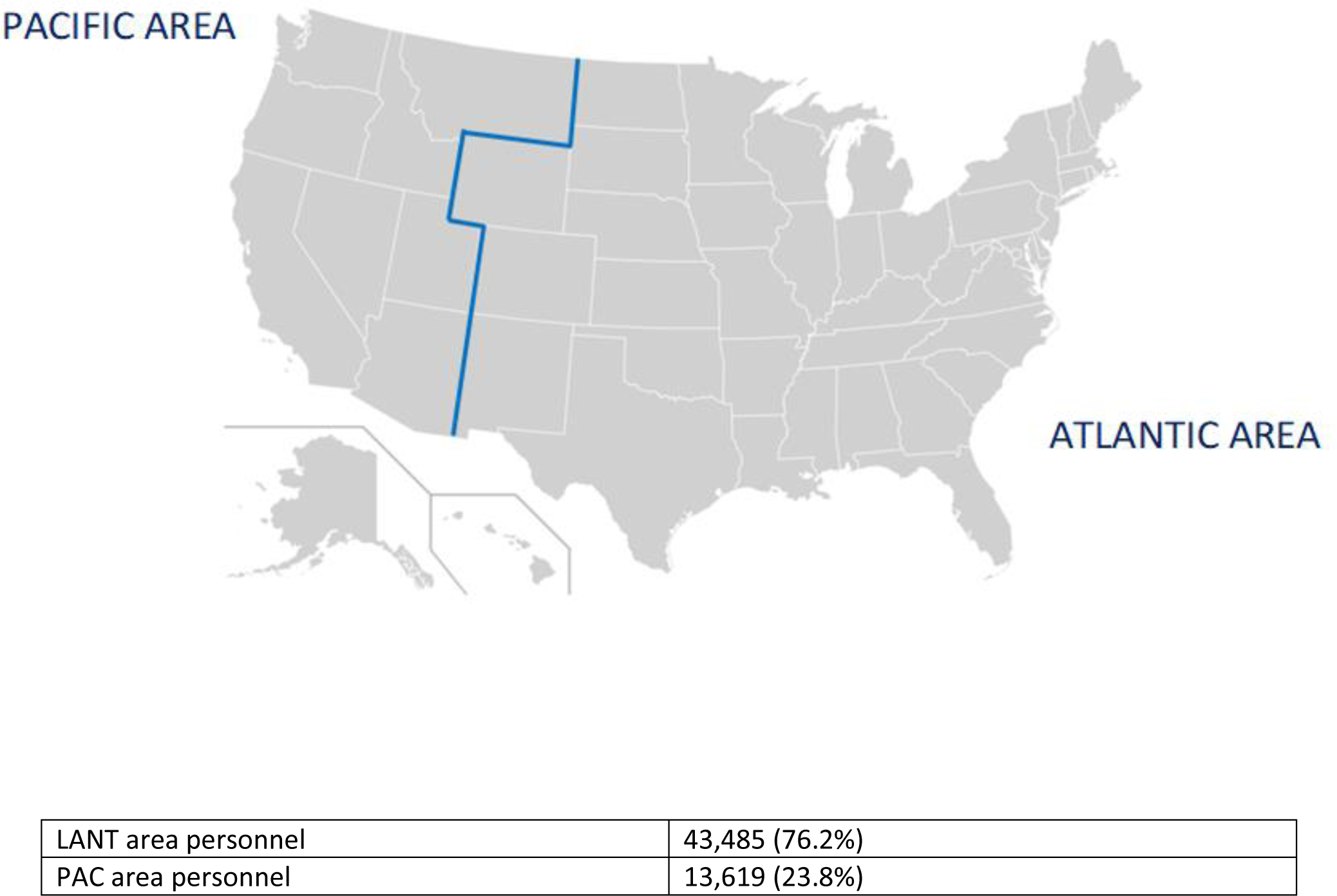
Atlantic (LANT) and Pacific (PAC) Areas within the U.S. Coast Guard (Source: CGPAAS)

Separately, we compared the rate of breakthrough infections seen by month between those reported as fully vaccinated with the Pfizer and Moderna vaccines. We considered a breakthrough infection to be any new report of a positive test for COVID-19 in an individual who had received two doses of either the Pfizer or Moderna vaccines, or a single dose of the J and J vaccine. For this analysis, we included CG members who had received the J and J vaccine.

The CG Institutional Review Board reviewed the investigation proposal and deemed the project exempt from the requirements of 45 CFR 46.

## Results

Overall VE among CG personnel in May 2021 was 82.6% and increased to 89.0% in June; declines were seen in each subsequent month of the study, reaching a nadir of 62.7% in August. CI were wider in May and June, reflecting fewer numbers of reported cases during the early period of the study. For June, the overall (unweighted) VE estimate was greater than either regional estimate because the PAC region had both a higher rate of full vaccination and fewer reported COVID-19 infections (Data Appendix). In both the LANT and PAC areas of the CG, declines in VE were noted in July and August. In the LANT area, VE declines were steeper than in the PAC area (Table 1). Table 2 shows age-stratified VE analyses over the course of the study. For all months except June, based on point estimates 17-24 y/o had the highest VE. Other notable findings were the wide variation in VE among the oldest age group: it was 100% in May but fell to −0.1 and 5.5% in July and August, respectively. Again decreasing width of the CI (and therefore increased precision of the estimates) was seen in July and August, due to increased numbers of reported cases (Data Appendix).

**Table 1:** Percent Vaccine Effectiveness and 95% CI Among Coast Guard Personnel Against Any New Report of COVID-19 Test Positivity, by Month and Geographic Area, 2021

|  | Overall | Pacific | Atlantic |
| --- | --- | --- | --- |
| May | 82.6 (78.2-86.2) | 78.5 (68.5-85.3) | 89.9 (80.3-94.8) |
| June | 89.0 (84.6-92.1) | 85.0 (-34.3-98.3) | 88.7 (83.2-92.5) |
| July | 68.9 (68.1-69.8) | 78.2 (76.4-79.9) | 66.2 (64.8-67.5) |
| August | 62.7 (62.4-63.1) | 70.3 (69.2-71.4) | 61.2 (60.6-61.7) |

**Table 2:** Percent Vaccine Effectiveness and 95% CI Among Coast Guard Personnel Against Any New Report of COVID-19 Test Positivity, by Month and Age, 2021

|  | 17-24 | 25-31 | 32-44 | ≥45 |
| --- | --- | --- | --- | --- |
| May | 93.6 (54.3-99.1) | 84.8 (70.1-92.3) | 57.6 (26.2-75.6) | 100.0 (NA) |
| June | 79.5 (-83.6-97.7) | 86.2 (72.7-93.0) | 93.8 (83.1-97.7) | NA |
| July | 87.5 (85.3-89.3) | 64.7 (61.4-67.6) | 71.5 (69.5-73.3) | -0.1 (-0.8-0.4) |
| August | 85.8 (85.1-86.4) | 80.9 (80.1-81.7) | 76.8 (76.2-77.3) | 5.5 (-20.6-25.9) |

During the four month period of this study, the rate of reported breakthrough infections in CG members did not differ between those fully vaccinated with Moderna or Pfizer vaccines (Table 3). For the first three months, the rate of breakthrough infections was not noticeably higher for J and J vaccine recipients; however in August the rate increased to 1.96%, approximately two and a half times the breakthrough rate seen with either two-dose vaccine. There were no COVID-19 related hospitalizations or deaths among fully vaccinated CG personnel during the study period, while eleven unvaccinated personnel were hospitalized.

**Table 3:** Number* and Percent of Newly Reported COVID-19 Infections Among Fully Vaccinated Personnel, by Month and Type of Vaccine Received

|  | Pfizer | Moderna | Johnson and Johnson |
| --- | --- | --- | --- |
| May | 6/25,107<br>(0.02%) | 3/10,946<br>(0.03%) | 0/992<br>(0.00%) |
| June | 3/26,236<br>(0.01%) | 2/11,389<br>(0.02%) | 0/1,478<br>(0.00%) |
| July | 61/26,639<br>(0.23%) | 23/11,538<br>(0.20%) | 5/1,759<br>(0.28%) |
| August | 216/27,706<br>(0.78%) | 97/11,960<br>(0.81%) | 46/2,342<br>(1.96%) |

\*New case reports in fully vaccinated/Total fully vaccinated based on mid-month data

## Discussion

Our findings showed VE decreasing from a high of near 90% to just under 63%, combined with continued protection against serious outcomes throughout the study period. Despite lack of generalizability due to the “healthy warrior” effect (Niebuhr et al 2011), study results are broadly in accord with other national analyses of vaccine effectiveness during the period when the Delta variant was becoming predominant (Rosenberg ES et al 2021, Herlihy R et al 2021, Nanduri S et al 2021). For example, in a study among frontline healthcare workers that also ran through August 2021, a decrease in VE from 91 to 66% was seen (Fowlkes A et al, 2021). A separate March-July 2021 analysis found 84% VE against hospitalization at 24 weeks following full 2-dose immunization (Tenforde MW et al. 2021). From April through mid-July 2021, a case-based report on persons ≥18 years old similarly found reduced VE for prevention of any infection, but continued high VE against severe COVID-19 related hospitalization and death (Scobie et al. 2021).

Over the course of this study, the circulation of SARS-CoV-2 within the U.S. underwent a transition from the Alpha variant being predominant during May 2021 (Paul P et al, 2021) through June during which the Delta variant represented approximately 30% of sequenced viruses, to over 80% by mid-July (S. Hadler, CDC, personal communication). In both the LANT and PAC areas of the CG, decreases in VE were noted in July and continued into August 2021. Given the general West to East spread of the Delta variant (Paul P et al 2021), we hypothesized that the PAC area might have seen decreases in VE sooner. While in May and June the PAC area had slightly lower estimated VE, the overlapping CI between PAC and LANT argue against this representing a real difference. One possible explanation for the steeper declines in VE in the LANT area in July and August is the area’s higher population and population density, which may have resulted in greater exposure to circulating virus among fully vaccinated personnel. Alternatively or additionally, the broad geographic groupings used for this analysis may have obscured more geographically discrete VE trends.

Our lowest VE estimates in this study occurred in the oldest age group during July and August. Recent serological and epidemiological studies support the idea that the immune response to and protection afforded by 2-dose mRNA vaccines may decrease with age (Bates TA et al 2021, Haas EJ et al 2021, Baden LR et al 2021). The finding during the final month of this study of lower breakthrough rates with 2-dose vaccines compared to single dose vaccines is consistent with recent U.S. findings of higher VE against serious outcomes and more robust antibody responses with 2-dose vaccines (Self WH et al, 2021).

The findings in this report are subject to at least four limitations. First, both COVID-19 case status and vaccination status are subject to underreporting. Verified vaccination status is subject to underreporting among civilians, which may have resulted in misclassification of some cases as not fully vaccinated.

Though this could have biased VE estimates upward, the number of civilians reporting cases was small. Second, there may have been biased reporting of cases. With recent attention to breakthrough infections (Brown CM et al, 2021), there may have been preferential reporting of test positivity among fully vaccinated individuals. Such a pattern would have biased our VE estimates downward. Third, the current analysis does not take into account time since vaccination. As the apparent need for additional doses in the COVID-19 vaccine schedule emerges (Tré-Hardy M et al 2021), the lower VE estimates seen later in this study may partially reflect waning immunity for those with earlier vaccination dates (Fowlkes A et al, 2021, Mizrahi B et al, 2021). Older age may have also correlated with time since vaccination, since early U.S. COVID-19 vaccination strategies (which were reflected in initial CG vaccination policy) were primarily based on increasing age as a risk factor (Dooling K et al, 2020). In other words, within CG age may serve as a proxy measure for time since vaccination. Finally, several wide CI seen early in the study due to relatively small numbers of reported cases (Data Appendix) indicate less precise estimates of VE than those seen later in the study when more cases were reported.

Both SARS-CoV-2 and other respiratory viruses including adenovirus have caused outbreaks among military personnel, especially in training settings (Chu VT et al, 2021) Given the benefits seen in this study and other broader measures of benefit such as lives saved seen with COVID-19 vaccines (Gupta S et al, 2021), policy and communications initiatives to increase vaccine uptake among military personnel may be warranted. On August 24 2021, the U.S. Secretary of Defense mandated COVID-19 vaccination for military personnel. The Commandant of the Coast Guard followed suit for CG Active Duty and Ready Reserve personnel on August 26.

A further implication of our study is that organizations that have ready access to diagnostic and immunization records may wish to monitor VE on an ongoing basis, as a way to facilitate both COVID-19 risk mitigation measures and more specific vaccine benefit and risk communication to their leadership and membership. This approach also maximizes use of available data, without the need for extensive statistical software or support. Another advantage of using VE as a key metric is that it is a population-based measure which reflects the experience of all members of an organization: the sick and the well, the vaccinated and the unvaccinated. Such population level measures may be easier to communicate than proportional morbidity analyses, where both the numerator and denominator require the onset and detection of illness.

In a geographically dispersed U.S. military population, VE against a newly reported COVID-19 diagnosis declined during a period of increasing circulation of the Delta variant. However by the end of the study period, the overall VE of just under 63 percent meets or exceeds influenza VE seen in years where circulating strains are well-matched to the vaccine (Treanor JJ et al 2012, Belongia E et al 2009). To date, there have been no hospital admissions due to COVID-19 among fully vaccinated Coast Guard personnel. In light of ongoing spread of the Delta variant and the possible emergence of other SARS-CoV-2 variants, to protect its workforce the Coast Guard plans ongoing monitoring of VE to include enhanced analyses such as time since vaccination, in addition to surveillance of other key public health and clinical measures related to COVID-19 disease.

## Data Availability

Data is available in the attached supplemental file.

## Disclaimer

The views expressed herein are the authors’, and should not be construed as the views of the U.S. Coast Guard, the Department of Defense (DoD), or the Department of Homeland Security (DHS).

Names of products or companies are included for identification purposes only and do not constitute any endorsement by the CG, DoD, or DHS.

## Acknowledgements

For technical consultation, we acknowledge Stephen Hadler MD and Paul Siegel MD, MPH from the Centers for Disease Control and Prevention, Tamara Ritsema PhD, MPH, PA-C from the George Washington University School of Medicine and Health Sciences, and Jonas Iskander from Harvard University. We also acknowledge all members of the Coast Guard COVID Crisis Action Team and COVID-19 Vaccine Incident Command for their service and sacrifice.

## Statement of Contributions

CAPT Iskander conceived of and drafted the manuscript. LT Frost and CDR Russell provided primary data analysis and critical review of the manuscript. CDR Ward, CDR Ahluwalia, and CAPT Steiner reviewed the manuscript for important intellectual content. RADM Thomas and CAPT Michaud reviewed the manuscript for intellectual content and provided materiel and leadership support.

## References

Marcus JE, Frankel DN, Pawlak MT, et al. COVID-19 Monitoring and Response Among U.S. Air Force Basic Military Trainees — Texas, March–April 2020. MMWR Morb Mortal Wkly Rep 2020;69:685–688.

Clifton GT, Pati R, Krammer F, et al. SARS-CoV-2 Infection Risk Among Active Duty Military Members Deployed to a Field Hospital — New York City, April 2020. MMWR Morb Mortal Wkly Rep 2021;70:308–311.

Yonker LM, Neilan AM, Bartsch Y, et al. Pediatric Severe Acute Respiratory Syndrome Coronavirus 2 (SARS-CoV-2): Clinical Presentation, Infectivity, and Immune Responses. J Pediatr. 2020 Dec;227:45-52.e5.

Rosenberg ES, Holtgrave DR, Dorabawila V, et al. New COVID-19 Cases and Hospitalizations Among Adults, by Vaccination Status — New York, May 3–July 25, 2021. MMWR Morb Mortal Wkly Rep. ePub: 18 August 2021.

Herlihy R, Bamberg W, Burakoff A, et al. Rapid Increase in Circulation of the SARS-CoV-2 B.1.617.2 (Delta) Variant — Mesa County, Colorado, April–June 2021. MMWR Morb Mortal Wkly Rep 2021;70:1084–1087.

Lopez Bernal J, Andrews N, Gower C, et al. Effectiveness of Covid-19 Vaccines against the B.1.617.2 (Delta) Variant. N Engl J Med. 2021 Aug 12;385(7):585–594.

Rubin D, Eisen M, Collins S, et al. SARS-CoV-2 Infection in Public School District Employees Following a District-Wide Vaccination Program — Philadelphia County, Pennsylvania, March 21–April 23, 2021. MMWR Morb Mortal Wkly Rep 2021;70:1040–1043.

Orenstein WA, Bernier RH, Dondero TJ, Hinman AR, Marks JS, Bart KJ, Sirotkin B (1985). Field evaluation of vaccine efficacy. Bull. World Health Organ. 63 (6): 1055–1068.

Niebuhr DW, Krampf RL, Mayo JA, Blandford CD, Levin LI, Cowan DN. Risk factors for disability retirement among healthy adults joining the U.S. Army. Mil Med. 2011 Feb;176(2):170–5.

Nanduri S, Pilishvili T, Derado G, et al. Effectiveness of Pfizer-BioNTech and Moderna Vaccines in Preventing SARS-CoV-2 Infection Among Nursing Home Residents Before and During Widespread Circulation of the SARS-CoV-2 B.1.617.2 (Delta) Variant — National Healthcare Safety Network, March 1–August 1, 2021. MMWR Morb Mortal Wkly Rep. ePub: 18 August 2021.

Fowlkes A, Gaglani M, Groover K, et al. Effectiveness of COVID-19 Vaccines in Preventing SARS-CoV-2 Infection Among Frontline Workers Before and During B.1.617.2 (Delta) Variant Predominance — Eight U.S. Locations, December 2020–August 2021. MMWR Morb Mortal Wkly Rep 2021;70:1167–1169.

Tenforde MW, Self WH, Naioti EA, et al. Sustained Effectiveness of Pfizer-BioNTech and Moderna Vaccines Against COVID-19 Associated Hospitalizations Among Adults — United States, March–July 2021. MMWR Morb Mortal Wkly Rep 2021;70:1156–1162

Scobie HM, Johnson AG, Suthar AB, et al. Monitoring Incidence of COVID-19 Cases, Hospitalizations, and Deaths, by Vaccination Status — 13 U.S. Jurisdictions, April 4–July 17, 2021. MMWR Morb Mortal Wkly Rep 2021;70:1284–1290.

Paul P, France AM, Aoki Y, et al. Genomic Surveillance for SARS-CoV-2 Variants Circulating in the United States, December 2020–May 2021. MMWR Morb Mortal Wkly Rep 2021;70:846–850.

Bates TA, Leier HC, Lyski ZL, et al. Age-Dependent Neutralization of SARS-CoV-2 and P.1 Variant by Vaccine Immune Serum Samples. JAMA. 2021 Jul 21;326(9):868–9.

Haas EJ, Angulo FJ, McLaughlin JM, et al. Impact and effectiveness of mRNA BNT162b2 vaccine against SARS-CoV-2 infections and COVID-19 cases, hospitalisations, and deaths following a nationwide vaccination campaign in Israel: an observational study using national surveillance data. Lancet. 2021;397 (10287):1819–1829.

Baden LR, El Sahly HM, Essink B, et al; COVE Study Group. Efficacy and safety of the mRNA-1273 SARS-CoV-2 vaccine. N Engl J Med. 2021;384(5):403–416

Self WH, Tenforde MW, Rhoads JP, et al. Comparative Effectiveness of Moderna, Pfizer-BioNTech, and Janssen (Johnson & Johnson) Vaccines in Preventing COVID-19 Hospitalizations Among Adults Without Immunocompromising Conditions — United States, March–August 2021. MMWR Morb Mortal Wkly Rep 2021;70:1337–1343.

Brown CM, Vostok J, Johnson H, et al. Outbreak of SARS-CoV-2 Infections, Including COVID-19 Vaccine Breakthrough Infections, Associated with Large Public Gatherings — Barnstable County, Massachusetts, July 2021. MMWR Morb Mortal Wkly Rep 2021;70:1059–1062.

Tré-Hardy M, Cupaiolo R, Wilmet A, et al. Six-month interim analysis of ongoing immunogenicity surveillance of the mRNA-1273 vaccine in healthcare workers: A third dose is expected. J Infect. 2021 Aug 23:S0163-4453(21)00433-3.

Mizrahi B, Lotan R, Kalkstein N, et al. Correlation of SARS-CoV-2 Breakthrough Infections to Time-from-vaccine;PreliminaryStudy. medRxiv 2021.07.29.21261317; doi: https://doi.org/10.1101/2021.07.29.21261317

Dooling K, Marin M, Wallace M, et al. The Advisory Committee on Immunization Practices’ Updated Interim Recommendation for Allocation of COVID-19 Vaccine — United States, December 2020. MMWR Morb Mortal Wkly Rep 2021;69:1657–1660.

Chu VT, Simon E, Lu X, et al. Outbreak of Acute Respiratory Illness Associated with Human Adenovirus Type 4 at the U.S. Coast Guard Academy, 2019. J Infect Dis. 2021 Jun 17:jiab322.

Gupta S, Cantor J, Simon KI, et al. Vaccinations Against COVID-19 May Have Averted Up To 140,000 Deaths In The United States. Health Aff (Millwood). 2021 Aug 18:101377hlthaff202100619.

Treanor JJ, Talbot HK, Ohmit SE, et al. Effectiveness of seasonal influenza vaccines in the United States during a season with circulation of all three vaccine strains. Clin Infect Dis. 2012 Oct;55(7):951–9.

Belongia EA, Kieke BA, Donahue JG, et al. Effectiveness of inactivated influenza vaccines varied substantially with antigenic match from the 2004-2005 season to the 2006-2007 season. J Infect Dis. 2009 Jan 15;199(2):159–67

