## Supplementary material for "Effectiveness of Vaccination against Reported SARS-CoV-2 Infection in United States Coast Guard Personnel between May and August 2021: A Time-Series Analysis": Data Appendix

Raw Data for Table 1

| May | | | | | |
|---|---|---|---|---|---|
| | PAC | LANT | | PAC | LANT |
| New case reports in fully vaccinated | 6 | 3 | New case reports in all other vaccine categories | 10 | 18 |
| Total fully vaccinated | 9994 | 27054 | Total in all other vaccine categories | 3583 | 16450 |
| Percentage | 0.06% | 0.01% | Percentage | 0.28% | 0.11% |

| June | | | | | |
|---|---|---|---|---|---|
| | PAC | LANT | | PAC | LANT |
| New case reports in fully vaccinated | 1 | 5 | New case reports in all other vaccine categories | 2 | 23 |
| Total fully vaccinated | 10441 | 28665 | Total in all other vaccine categories | 3136 | 14839 |
| Percentage | 0.01% | 0.02% | Percentage | 0.06% | 0.15% |

| July | | | | | |
|---|---|---|---|---|---|
| | PAC | LANT | | PAC | LANT |
| New case reports in fully vaccinated | 30 | 59 | New case reports in all other vaccine categories | 39 | 84 |
| Total fully vaccinated | 10582 | 29357 | Total in all other vaccine categories | 2995 | 14147 |
| Percentage | 0.28% | 0.20% | Percentage | 1.30% | 0.59% |

| August | | | | | |
|---|---|---|---|---|---|
| | PAC | LANT | | PAC | LANT |
| New case reports in fully vaccinated | 83 | 186 | New case reports in all other vaccine categories | 67 | 192 |
| Total fully vaccinated | 10953 | 31058 | Total in all other vaccine categories | 2624 | 12446 |
| Percentage | 0.76% | 0.60% | Percentage | 2.55% | 1.54% |

Raw Data for Table 2

| | | | | | | | | | |
|---|---|---|---|---|---|---|---|---|---|
| May | | | | | | | | | |
| | 17-24 | 25-31 | 32-44 | ≥45 | | 17-24 | 25-31 | 32-44 | ≥45 |
| New case reports in fully vaccinated | 1 | 3 | 5 | 0 | New case reports in all other vaccine categories | 9 | 11 | 5 | 2 |
| Total fully vaccinated | 7014 | 8008 | 15521 | 5445 | Total in all other vaccine categories | 4012 | 4465 | 6583 | 3035 |
| Percentage | 0.01% | 0.04% | 0.03% | 0.00% | Percentage | 0.22% | 0.25% | 0.08% | 0.07% |
| June | | | | | | | | | |
| | 17-24 | 25-31 | 32-44 | ≥45 | | 17-24 | 25-31 | 32-44 | ≥45 |
| New case reports in fully vaccinated | 1 | 3 | 2 | 0 | New case reports in all other vaccine categories | 2 | 10 | 12 | 0 |
| Total fully vaccinated | 7817 | 8546 | 16079 | 5525 | Total in all other vaccine categories | 3209 | 3927 | 6025 | 2955 |
| Percentage | 0.01% | 0.04% | 0.01% | 0.00% | Percentage | 0.06% | 0.25% | 0.20% | 0.00% |
| July | | | | | | | | | |
| | 17-24 | 25-31 | 32-44 | ≥45 | | 17-24 | 25-31 | 32-44 | ≥45 |
| New case reports in fully vaccinated | 13 | 29 | 38 | 8 | New case reports in all other vaccine categories | 37 | 35 | 47 | 4 |
| Total fully vaccinated | 8128 | 8744 | 16339 | 5555 | Total in all other vaccine categories | 2898 | 3729 | 5765 | 2925 |
| Percentage | 0.16% | 0.33% | 0.23% | 0.14% | Percentage | 1.28% | 0.94% | 0.82% | 0.14% |
| August | | | | | | | | | |
| | 17-24 | 25-31 | 32-44 | ≥45 | | 17-24 | 25-31 | 32-44 | ≥45 |
| New case reports in fully vaccinated | 54 | 60 | 134 | 18 | New case reports in all other vaccine categories | 61 | 66 | 114 | 9 |
| Total fully vaccinated | 9500 | 10310 | 18454 | 5758 | Total in all other vaccine categories | 1526 | 2163 | 3650 | 2722 |
| Percentage | 0.57% | 0.58% | 0.73% | 0.31% | Percentage | 4.00% | 3.05% | 3.12% | 0.33% |
